# Evaluation of Large Language Model-Generated Patient Information for Communicating Radiation Risk

**DOI:** 10.1101/2025.07.23.25332093

**Authors:** Alice Gutowski, Daniel Carrion, Mohamed Khaldoun Badawy

## Abstract

**Background:** Large language models are increasingly used to generate patient information in healthcare. However, their ability to communicate complex topics, such as the risks associated with radiation from medical imaging, remains unclear.

**Purpose:** This study evaluated the quality, relevance, and readability of patient information generated by large language models for communicating radiation risks associated with computed tomography and interventional cardiology procedures.

**Methods:** Five large language models were prompted to generate patient information for two clinical scenarios: computed tomography and interventional cardiology. The information was assessed by medical physicists, radiographers, and health literacy specialists using a structured survey containing rating scales and free-text feedback. Statistical analyses included tests for normality, group comparisons using non-parametric methods, and thematic analysis of qualitative responses.

**Results:** Twelve healthcare professionals participated. Significant differences were identified among professional groups in their scoring of readability, language suitability, and tone, particularly for higher-risk procedures. Health literacy specialists reported significant differences between large language models across most criteria, while medical physicists and radiographers identified fewer differences. Qualitative feedback revealed variability in how well the models balanced technical accuracy with accessible language, with some including inaccurate or irrelevant information.

**Conclusion:** Large language models show potential in supporting the development of patient information for radiation risk communication; however, substantial variability remains in the quality and appropriateness of the content. Multidisciplinary review is essential, and sole reliance on large language model-generated materials is not recommended. Further research involving patient evaluation is required to assess the real-world impact of these tools in clinical settings.

## Introduction

The integration of artificial intelligence (AI) into healthcare is progressing rapidly, with large language models (LLMs) increasingly used to generate patient-facing information [1]. These tools offer the potential to enhance understanding, reduce health literacy barriers, and support shared decision-making [2]. While many studies have assessed the accuracy, readability, and clinical relevance of LLM-generated information, this research has predominantly focused on general medical conditions and patient concerns, treatment overviews, or clinical correspondence [3] [4] [5] [6] [7] [8] [9] [10] [11].

Radiology and medical imaging present a unique context for patient communication, particularly regarding exposure to ionising radiation. Patient anxiety surrounding diagnostic or interventional imaging is well documented and may influence decision-making or compliance with recommended procedures [12] [13]. Effective risk communication is, therefore, crucial to support informed consent; however, existing approaches vary considerably [14]. In typical clinical practice, for lower-risk examinations, such as computed tomography (CT) scans, radiation risks are often communicated through posters or leaflets displayed in clinical areas. In contrast, higher-risk procedures, such as interventional cardiology, require more detailed verbal discussions and written information to explain potential deterministic effects.

Despite the growing use of LLMs to generate patient education materials [9] [10] [11], no studies to date have specifically evaluated their ability to communicate radiation risks to patients. Radiation risk communication is inherently complex, requiring accurate, accessible, and appropriately balanced information to support informed decision-making [15] [16].

This study aimed to assess the quality, relevance, and readability of LLM-generated patient information on radiation risks for both low- and high-risk imaging procedures. The findings will inform the potential role of LLMs in supporting radiation risk communication within clinical practice.

## Methods

### Study Design

This study evaluated the quality and relevance of patient information generated by LLMs for communicating radiation risks associated with medical imaging procedures. Five LLMs were selected for assessment based on their widespread use in healthcare research. These included: Copilot (GPT-4-turbo, accessed 2025-03-26), ChatGPT (4o-2025-01-29), Claude (3.5 Sonnet-2024-10-22), Llama (3.3-70b-instruct), and Mistral (small-24b-instruct-2501). These were chosen based on common models chosen in existing research in this area [3] [4] [5] [6] [7] [8].

Two clinical scenarios were selected to represent procedures with differing levels of radiation risk and corresponding information requirements: a CT scan and an interventional cardiology procedure. The CT scan represents a standard, lower-risk examination where brief patient information is typically provided. In contrast, interventional cardiology procedures carry a higher risk of deterministic effects, requiring more detailed risk communication.

To ensure consistency, a standardised prompt was developed and piloted for each scenario. These prompts were refined through iterative testing with two medical physicists to optimise clarity and relevance. The final prompts were:

- **Prompt 1:** *Generate educational information to provide to patients before undergoing an interventional cardiology procedure. Include a brief, accessible overview of the radiation risks to support informed decision-making. The information should be appropriate for patients with diverse health literacy, age, and cultural backgrounds*.
- **Prompt 2:** *Generate educational information to provide to patients before undergoing a CT scan. Include a brief, accessible overview of the radiation risks to support informed decision-making. The information should be appropriate for patients with diverse health literacy, age, and cultural backgrounds*.

All LLMs, except CoPilot, were accessed simultaneously using the Chatbot Arena platform (https://chat.lmsys.org/), which allows for unbiased comparison of responses from multiple models within the same interface and under identical conditions. CoPilot was accessed separately through the institutional licence, as this model is integrated into the organisation’s IT environment. This ensured the local, deployed version was evaluated alongside publicly accessible models.

### Survey Design and Data Collection

The generated information sheets were anonymised and compiled for assessment. A web-based survey was developed using Google Forms to evaluate the quality, relevance, and readability of each information sheet. The survey was reviewed for content validity by the two authors, both medical physicists with over 10 and 5 years of professional experience, respectively, before distribution.

Three groups of healthcare professionals were invited to participate: medical physicists, radiographers, and health literacy specialists. These groups were selected to provide complementary perspectives on the technical accuracy, clinical relevance, and readability of the generated information. Participants were recruited through internal departmental emails at the study site, with voluntary and anonymous participation.

The survey contained the following components:

#### Demographic Information

1. Job role (medical physicist, radiographer, health literacy specialist). **Evaluation Criteria:**For each information sheet, participants rated on a 5-point Likert scale (1 = Poor, 5 = Excellent):
2. Accuracy of procedural information.
3. Accuracy of radiation risk information.
4. Relevance of content to the examination and associated risks.
5. Comprehensiveness of radiation risk discussion.
6. Balance of risk and benefit conveyed.
7. Readability and ease of understanding.
8. Language appropriateness for diverse patient groups.
9. Tone suitability for risk communication. **Qualitative Feedback :**Free-text fields for anticipated patient response and additional comments.

### Data analysis

All data were analysed using Python version 3.11.7 with the following libraries: pandas (2.1.4), numpy (v1.26.4), scipy (v1.11.4), and matplotlib (v3.8.0). Statistical significance was set at p < 0.05.

Data normality was assessed using the Shapiro-Wilk test. As the data were not normally distributed, non-parametric tests were applied. Median scores and interquartile ranges were calculated for each model and professional group.

The Kruskal-Wallis test was used to assess differences in responses between professional groups for each evaluation criterion. Within-group comparisons of LLM performance were conducted using the Friedman test for related samples. Where significant differences were identified, post hoc pairwise comparisons with Bonferroni correction were applied.

Free-text responses underwent thematic analysis following the Braun and Clarke framework [17]. Themes were not pre-defined but emerged through inductive analysis of the qualitative data, consistent with the Braun and Clarke framework. The two authors independently coded the data, with discrepancies resolved through discussion to enhance the reliability of the results.

## Results

A total of 12 respondents completed the survey, comprising four medical physicists, four radiographers, and four health literacy specialists.

Table 1 presents the median overall score and interquartile range for each LLM, combining results from both clinical scenarios. Overall, ChatGPT, Llama, and Mistral achieved higher median scores compared to Claude and Copilot.

**Table 1.** Median (interquartile range) overall score for each LLM, by professional group and combined.

| **Model** | **Medical Physicists** | **Radiographers** | **Health Literacy** | **Overall** |
| --- | --- | --- | --- | --- |
| ChatGPT (4o-latest-20250129) (1) | 4.0 (4.0 - 5.0) | 4.0 (3.0 - 4.0) | 3.0 (2.0 - 4.0) | 4.0 (3.0 - 4.0) |
| Llama (3.3-70b-instruct) (2) | 4.0 (3.0 - 5.0) | 4.0 (3.0 - 4.0) | 3.0 (2.8 - 4.0) | 4.0 (3.0 - 4.0) |
| Claude (3.5 Sonnet-20241022) (3) | 3.5 (3.0 - 4.0) | 4.0 (3.0 - 4.0) | 3.0 (2.8 - 4.0) | 3.0 (3.0 - 4.0) |
| Mistral (small-24b-instruct-2501) (4) | 4.0 (3.0 - 4.0) | 3.0 (3.0 - 4.0) | 4.0 (3.0 - 4.0) | 4.0 (3.0 - 4.0) |
| Copilot (GPT-4-turbo) (5) | 4.0 (3.0 - 4.0) | 3.0 (3.0 - 4.0) | 2.0 (2.0 - 2.0) | 3.0 (2.0 - 4.0) |

Figures 1a–1f illustrate the distribution of scores for each evaluation criterion, stratified by professional group and clinical scenario.

**Fig 1(a)-(f).**
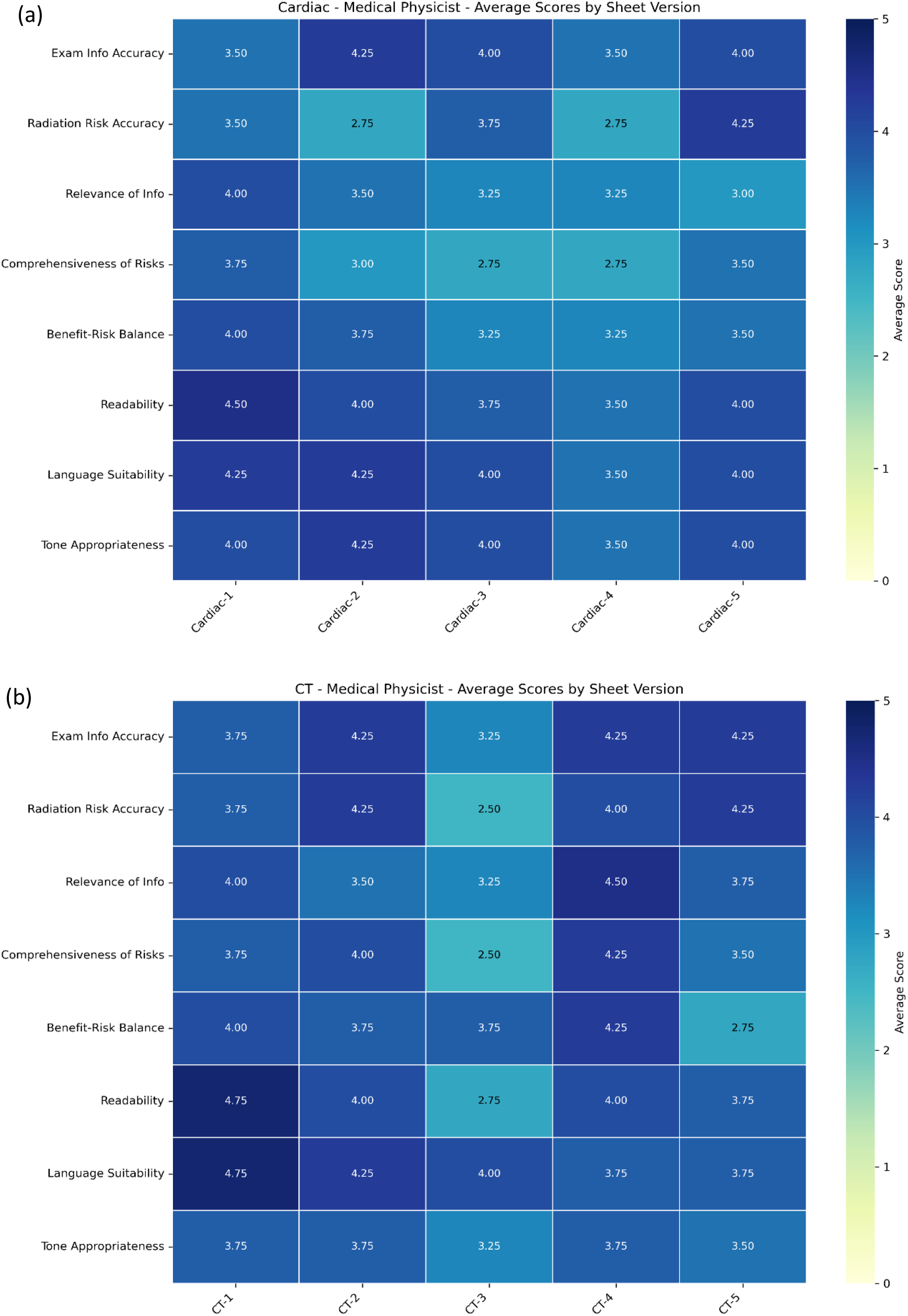
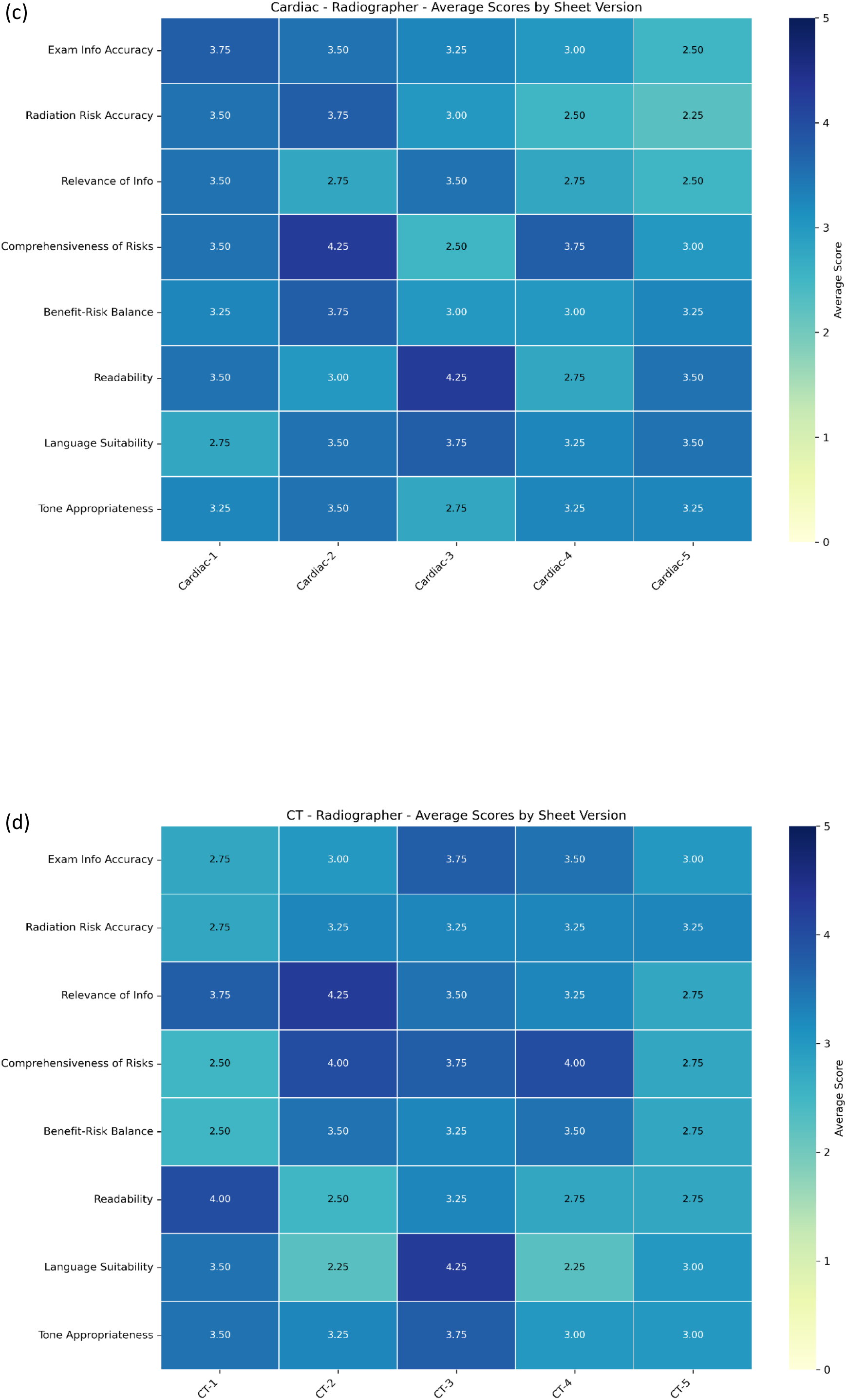
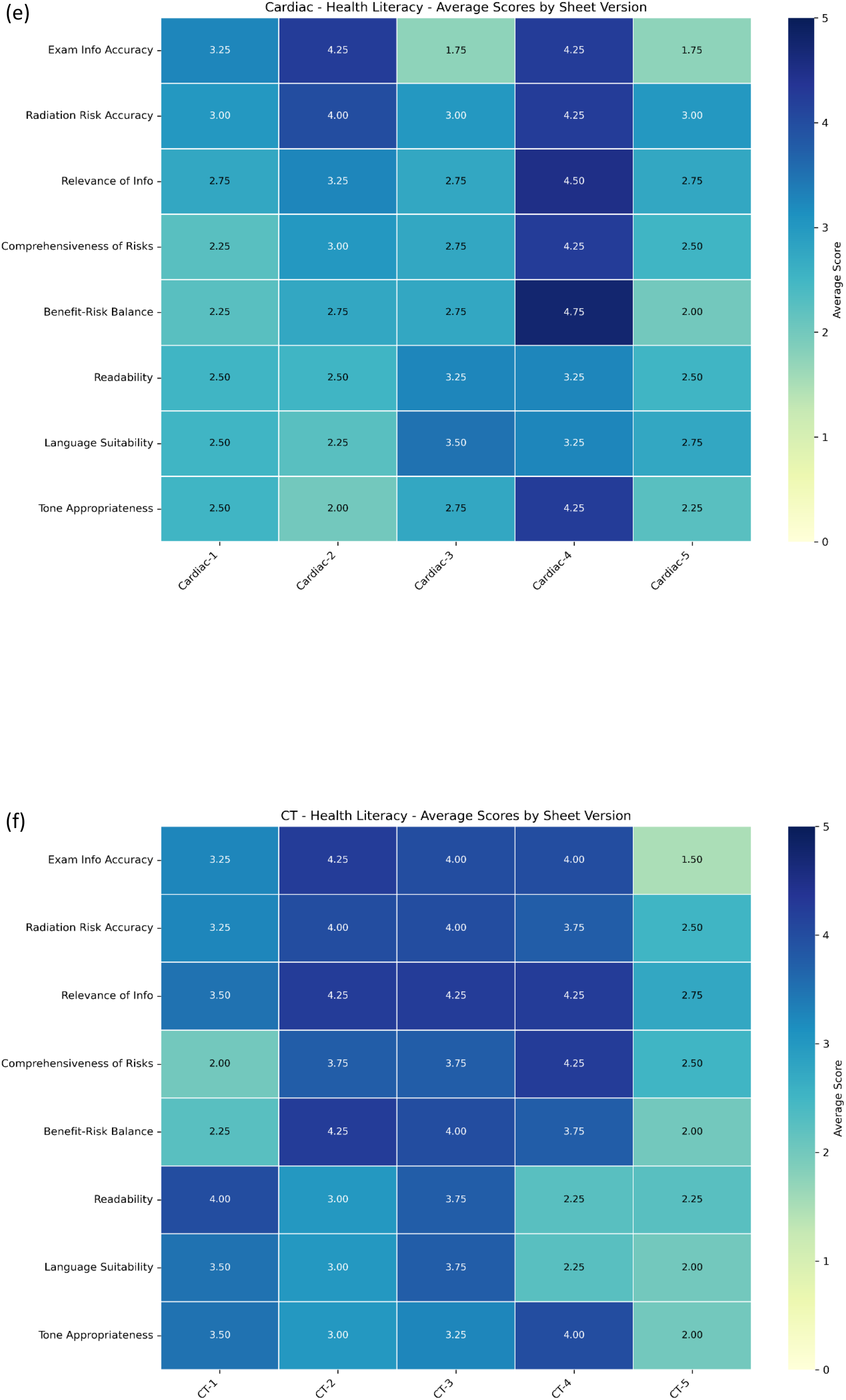
Heatmaps of the average score across all participants across all numerical evaluation criteria, for medical physicists, radiographers and health literacy, split by information sheet modality

The Kruskal–Wallis test was conducted to examine differences in evaluation scores between professional groups. For the CT scenario, significant differences were observed in readability, *p* = .049 and language appropriateness, *p* = .002. In the interventional cardiology scenario, significant differences were identified in the accuracy of information, *p* = .012; comprehensiveness of the radiation risk discussion, *p* = .043; readability, *p* = .001; language appropriateness, *p* <.001; and tone suitability, *p* = .002.

The Friedman test was used to assess whether significant differences existed in LLM performance within each professional group. Among medical physicists, no statistically significant differences were found between the LLMs for any evaluation criterion in either clinical scenario. Similarly, no significant differences were observed among radiographers for the interventional cardiology scenario. However, for the CT scenario, a significant difference was found in the “Relevance of information” criterion, *p* = .023. Among health literacy specialists, significant differences between LLMs were observed across most evaluation criteria for both scenarios, *p* = .050. Evaluation criteria without a significant difference included “Radiation risk information” for both the interventional cardiology scenario, *p* = .128, and CT, *p* = .161, and “Readability” for interventional cardiology, *p* = .114

When responses from all professional groups were combined, the Friedman test revealed statistically significant differences between LLMs for several evaluation criteria. These included accuracy of information, *p* = .013; relevance of information, *p* = .001; comprehensiveness of the radiation risk discussion, *p* = .001; balance of risk and benefit conveyed, *p* = .007; readability, *p* = .020; and language appropriateness, *p* <.001.

Thematic analysis of free-text responses identified recurring strengths and weaknesses for each LLM, summarised in Table 2. The identified themes were developed through inductive coding of participant responses, reflecting recurring issues related to accuracy and relevance of information, risk communication and emotional impact, language complexity and clarity, and patient engagement.

**Table 2.** Summary of the common themes and phrases identified in the free-text questions, categorised by model and job role. Each category is named, and the associated theme given, where T1 represents accuracy and relevance of information, T2 represents risk communication and emotional impact, T3 represents language complexity and clarity and T4 represents patient engagement.

| **Model** | **Medical Physicist** | **Radiographer** | **Health Literacy** |
| --- | --- | --- | --- |
| ChatGPT (4o-latest-20250129) (1) | Factual inaccuracies (T1)<br>Overemphasis on risk (T2) | Missing critical details (T1)<br>Appropriate literacy level (T3)<br>Supports patient questioning (T4) | Missing critical details (T1)<br>Appropriate literacy level (T3)<br>Supports patient questioning (T4)<br>Unmet information needs (T4) |
| Llama (3.3-70b-instruct) (2) | Factual inaccuracies (T1)<br>Alarmist tone (T2)<br>Overemphasis on risk (T2)<br>Overly complex language (T3) | Alarmist tone (T2)<br>Overly complex language (T3) | Missing critical details (T1)<br>Alarmist tone (T2)<br>Overemphasis on risk (T2)<br>Overly complex language (T3) |
| Claude (3.5 Sonnet-20241022) (3) | Missing critical details (T1)<br>Over-simplification (T3) | Missing critical details (T1)<br>Appropriate literacy level (T3)<br>Unmet information needs (T4) | Appropriate literacy level (T3)<br>Unmet information needs (T4) |
| Mistral (small-24b-instruct-2501) (4) | Irrelevant information (T1)<br>Overemphasis on risk (T2)<br>Unmet information needs (T4) | Irrelevant information (T1)<br>Overly complex language (T3) | Overly complex language (T3)<br>Supports patient questioning (T4) |
| Copilot (GPT-4-turbo) (5) | Irrelevant information (T1)<br>Factual inaccuracies (T1)<br>Overemphasis on risk (T2)<br>Unmet information needs (T4) | Irrelevant information (T1)<br>Factual inaccuracies (T1)<br>Overemphasis on risk (T2) | Missing critical details (T1)<br>Overly complex language (T3)<br>Unmet information needs (T4) |

## Discussion

This study evaluated the quality, relevance, and readability of LLM-generated patient information for communicating radiation risks in medical imaging. The findings demonstrate that while LLMs can produce generally acceptable information, significant variability exists between models and across different professional perspectives. Notably, no single LLM consistently outperformed others across all evaluation criteria or professional groups.

Differences in scoring between professional groups were most pronounced in areas relating to readability, language appropriateness, and tone. Although there are no existing studies directly comparing these groups, previous research does demonstrate that those with expertise in or high levels of health literacy may apply more stringent standards when evaluating patient information [18]. The health literacy specialists in this study identified significant differences between LLMs across most criteria, suggesting that LLM-generated information may not yet consistently meet best practice standards for accessible, patient-centred communication.

In contrast, medical physicists and radiographers demonstrated fewer significant differences between LLMs, with their assessments appearing to focus primarily on the presence of key technical information. This difference in evaluation emphasis highlights the need for a multidisciplinary approach when assessing or implementing LLM-generated patient materials. While technical accuracy is essential, appropriate language, readability, and tone are equally important to ensure information is accessible, reduce patient anxiety, and support informed decision-making [19].

Thematic analysis of qualitative feedback further supported these findings. LLMs varied in their ability to balance technical content with accessible language. For example, ChatGPT was generally considered easy to understand but lacked sufficient detail in some areas, while Llama provided more detailed information but used complex or potentially alarming language. Several models included inaccurate or irrelevant information, such as references to protective equipment or genetic risks, which may confuse or unnecessarily concern patients.

These results are consistent with recent studies assessing LLMs in other healthcare domains, which have similarly reported variability in accuracy, relevance, and appropriateness of LLM-generated patient information [3] [4] [5] [6] [7] [8] [9] [10] [11]. Importantly, this study extends those findings to the underexplored area of radiation risk communication, where the complexity of the topic and potential for patient anxiety underscore the need for clear, accurate, and carefully worded information.

This study has several limitations. First, the small sample size (n = 12 participants) limits the statistical power of the study, particularly in detecting more subtle differences between models. As such, these results should be interpreted with caution and may not be fully generalisable to broader clinical or patient populations. Second, LLMs are evolving rapidly; the specific model versions assessed may have evolved by the time of publication. Third, only healthcare professionals assessed the generated information. While these experts are well-positioned to evaluate the accuracy and appropriateness of LLM-generated materials, the actual impact on patient understanding, anxiety, and decision-making can only be determined through direct patient feedback.

These findings suggest that while LLMs may assist in drafting or refining radiation risk information, sole reliance on LLM-generated content is not yet recommended. A multidisciplinary review process, involving technical experts and health literacy specialists, remains essential to ensure accuracy, accessibility, and appropriateness. Future research should include patient-centred evaluations of LLM-generated materials to assess their real-world impact on understanding, satisfaction, and informed consent.

Additionally, as LLM technology continues to evolve, ongoing assessment will be required to monitor improvements in model performance and suitability for use in radiation risk communication.

## Conclusions

This study evaluated the ability of large language models (LLMs) to generate patient information for communicating radiation risks associated with medical imaging procedures. While LLMs demonstrated potential in producing relevant and generally understandable content, considerable variability was observed in the quality, accuracy, and readability of the information generated. No single LLM consistently performed well across all evaluation criteria or professional groups, and significant differences were identified in how technical experts, such as medical physicists and radiographers, and health literacy specialists assessed the materials. These findings underscore the importance of a multidisciplinary approach to evaluating LLM-generated patient information, particularly in the context of complex topics such as radiation risk. Although LLMs may serve as a useful tool to support the development of patient-facing materials, their use in isolation is not recommended. Careful human review, with input from both technical and health literacy experts, remains essential to ensure that information provided to patients is accurate, balanced, and accessible. As LLM technology continues to evolve, ongoing evaluation will be necessary to monitor improvements in model performance and suitability for clinical communication. Future research should incorporate direct patient feedback through usability testing, comprehension assessments, or anxiety measures to determine the real-world effectiveness of these materials.

## Data Availability

All data produced in the present study are available upon reasonable request to the authors

## Statements and Declarations

### Author Contributions

All authors contributed to the study conception and design. AG completed survey design, data collection and analysis. MB and AG made significant contributions to the draft writeup. All authors read and approved the final manuscript.

### Ethics Approval

No patient or sensitive data was collected for this project; hence an ethics review was not required.

### Informed Consent

Not applicable.

### Consent to Participate

Not applicable.

### Consent to Publish

Not applicable.

